## Supplemental Figures for "The Extra-Islet Pancreas Supports Autoimmunity in Human Type 1 Diabetes"

### Supplemental Information

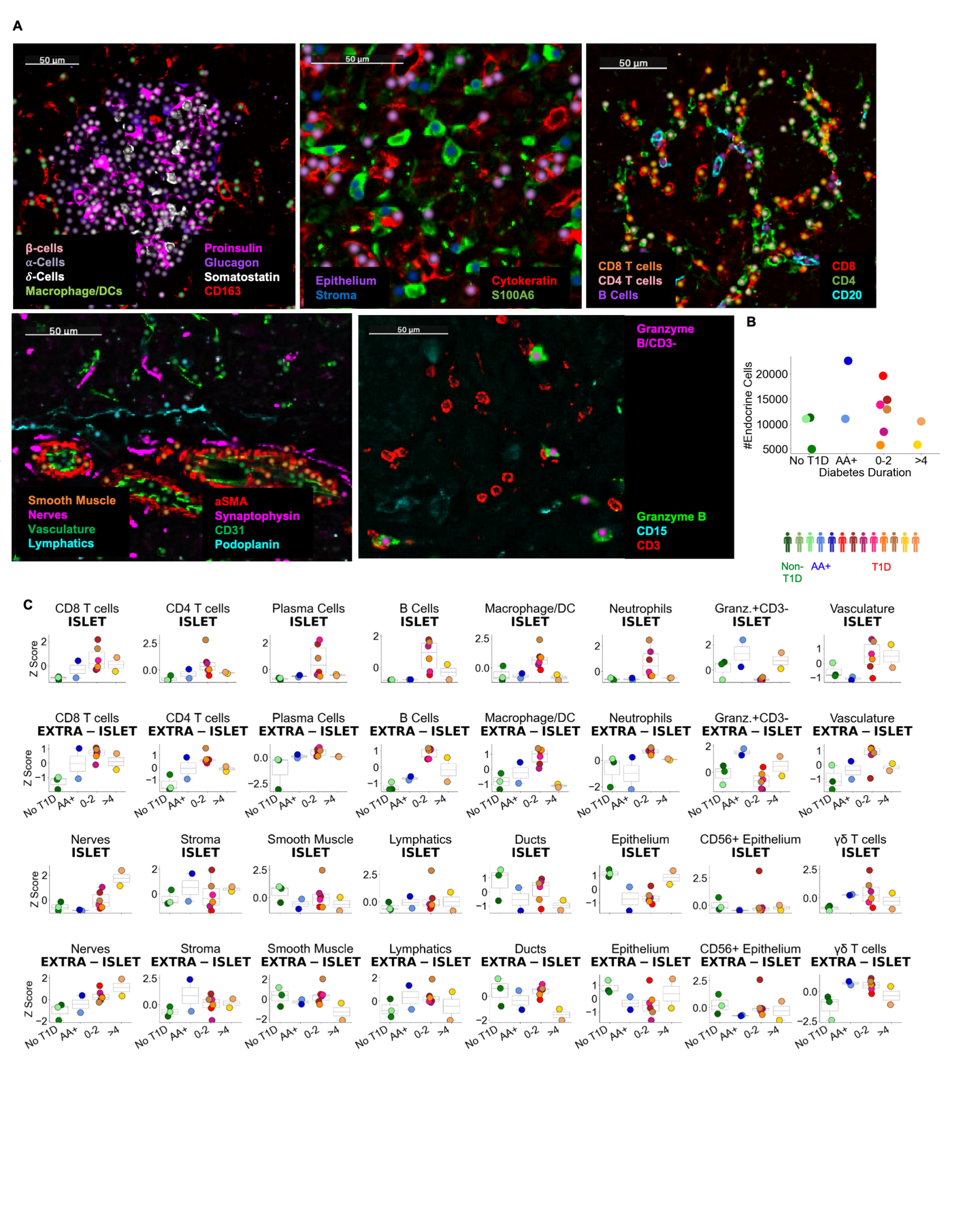

### Supplemental Figure 1 Related to Figure 1.

Supplemental Figure 1.A Validation of cell annotations. Dots were overlaid on images of characteristic markers colored by cell type. In each panel, the cell types and their corresponding colors are indicated and the markers and their corresponding colors are specified.

Supplemental Figure 1.B The total number of endocrine cells measured in each donor.

Supplemental Figure 1.C Changes in cellular abundance in Islet (top) and extra-islet (bottom) regions. The Y-axis corresponds to the number of the given cell type / number of endocrine cells in the top row and the number of a given cell type / number of acinar cells in the bottom row in each donor. These frequencies were then z-normalized across donors.

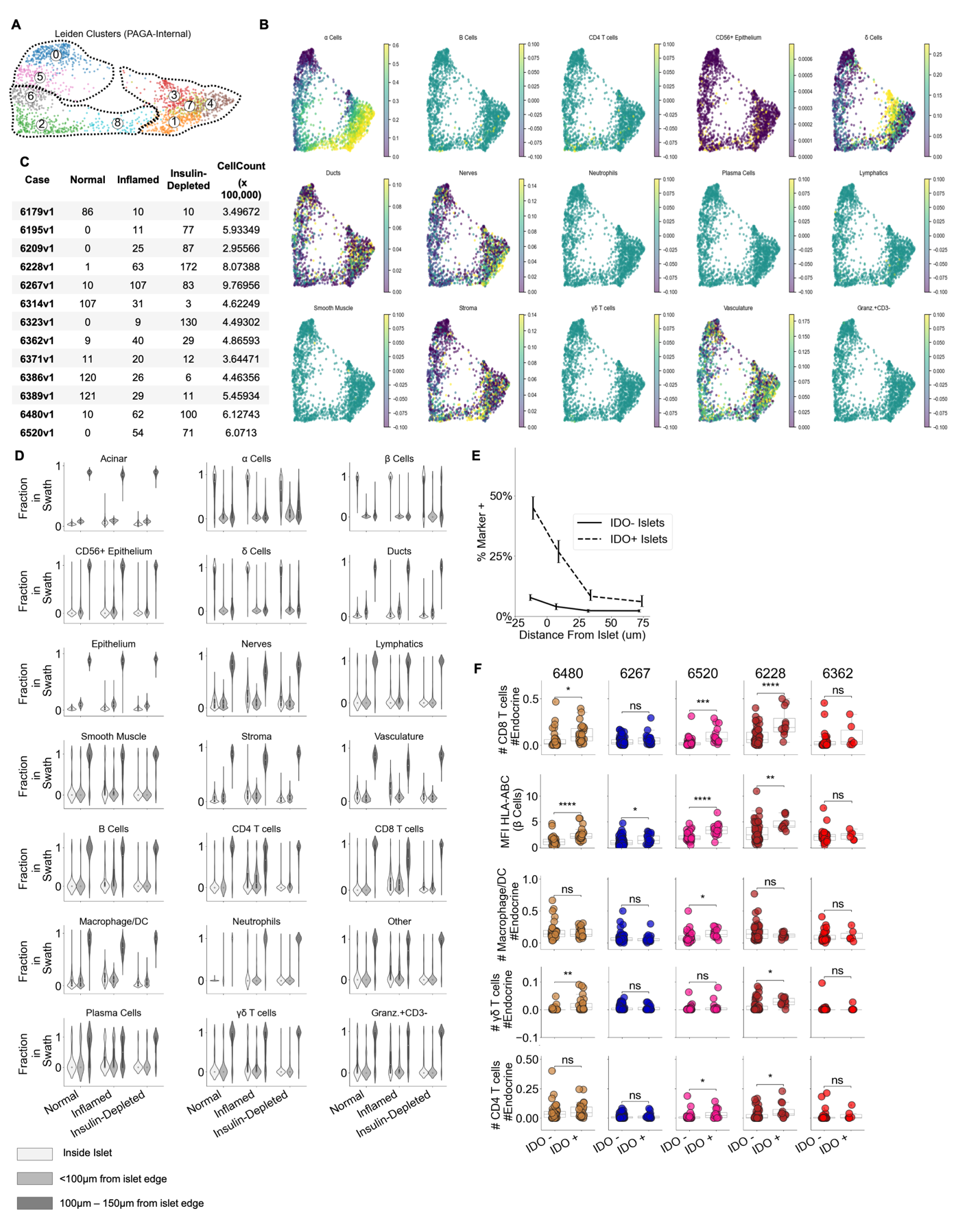

### Supplemental Figure 2 Related to Figure 2.

Supplemental Figure 2.A Leiden clustering computed by PAGA algorithm internally. Clusters 0 and 5 were assigned to the ‘Healthy’ group. Clusters 6, 2, and 8 were assigned to the “Inflamed” group. Clusters 1,3,7, and 4 were assigned to the “Insulin-Depleted” group.

Supplemental Figure 2.B The density of other cell types per islet across pseudotime. Same as Figure 2.E.

Supplemental Figure 2.C The number of islets of each stage of pseudotime and the total number of cells per case.

Supplemental Figure 2.D For each cell type, the frequency of that cell type inside islets, within 100µm of the islet edge, and 100µm-150µm from the islet edge was quantified.

Supplemental Figure 2.E Frequency of IDO on vasculature at different distances from islets. Dashed line indicates the frequency in and around islets where IDO^+^ was detected in islet vasculature (n=84) as in Figure 2.H. Solid line indicates the frequency in and around Inflamed Islets in which IDO was absent in islet vasculature (n=267). Error bars indicate 95% confidence intervals obtained by iteratively calculating the marker frequency in re-sampled islets with replacement (n=200)

Supplemental Figure 2.F CD8^+^T cell, Macrophage, γ/δ T cell, and CD4^+^T cell abundance in IDO^+^ and IDO^-^ islets. Only the mean expression of HLA-ABC in β-cells was measured in each islet. The other parameters measure the frequency of the respective cell type relative to the total number of endocrine cells. Same as Figure 2.I. Asterisks in figure indicate significance within each donor (Satterthwaites’s method lmerTest R package).

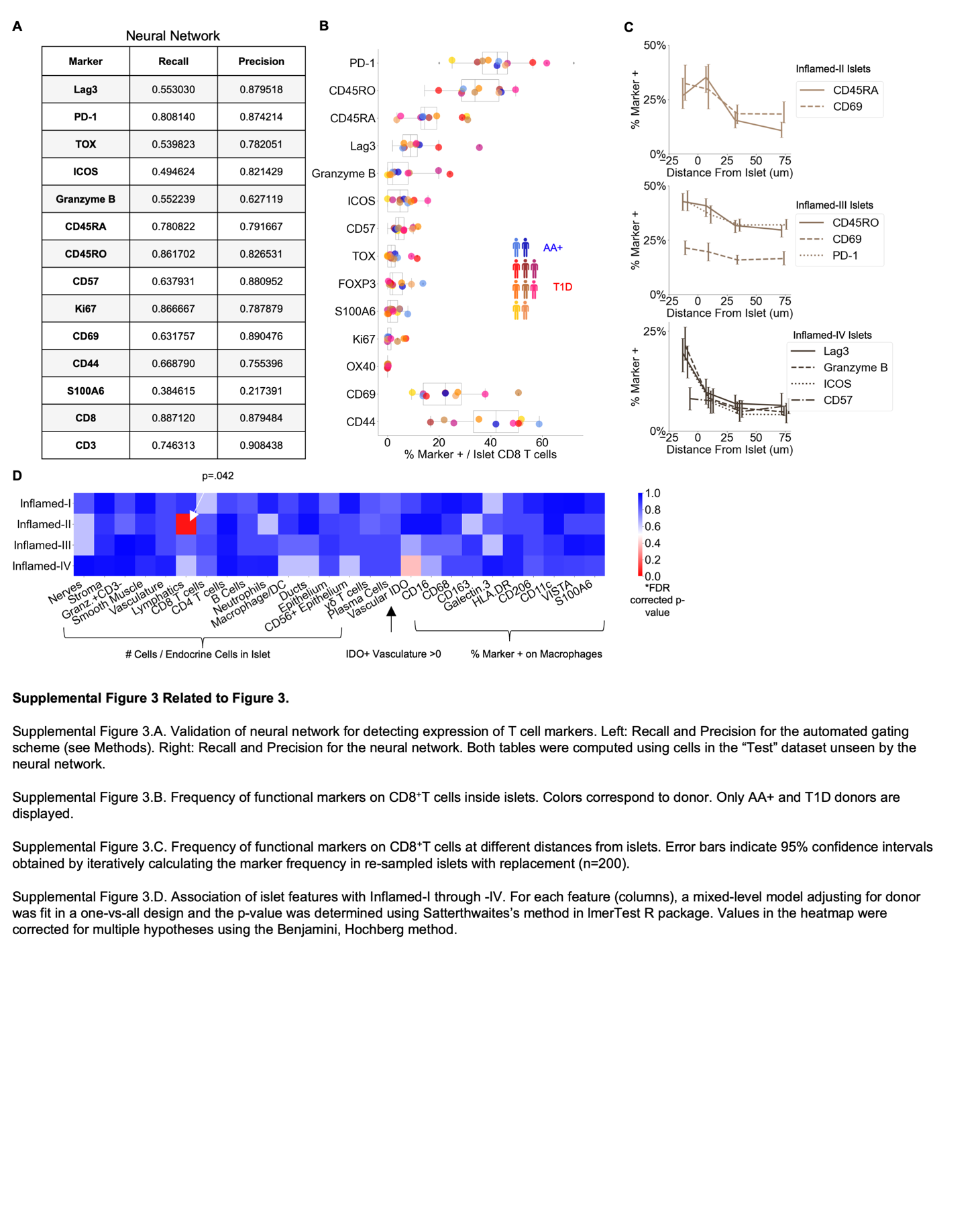

### Supplemental Figure 3 Related to Figure 3

Supplemental Figure 3.A Validation of neural network for detecting expression of T cell markers. Recall and Precision for the neural network. Left: Recall and Precision for the automated gating scheme (see Methods). Right: Recall and Precision for the neural network. Both tables were computed using cells in the “Test” dataset unseen by the neural network.

Supplemental Figure 3.B Frequency of functional markers on CD8^+^T cells inside islets. Colors correspond to donor. Only AA+ and T1D donors are displayed.

Supplemental Figure 3.C Frequency of functional markers on CD8^+^T cells at different distances from islets. Error bars indicate 95% confidence intervals obtained by iteratively calculating the marker frequency in re-sampled islets with replacement (n=200).

Supplemental Figure 3.D Association of islet features with Inflamed-I through -IV. For each feature (columns), a mixed-level model adjusting for donor was fit in a one-vs-all design and the p-value was determined using Satterthwaites’s method in lmerTest R package. Values in the heatmap were corrected for multiple hypotheses using the Benjamini, Hochberg method.

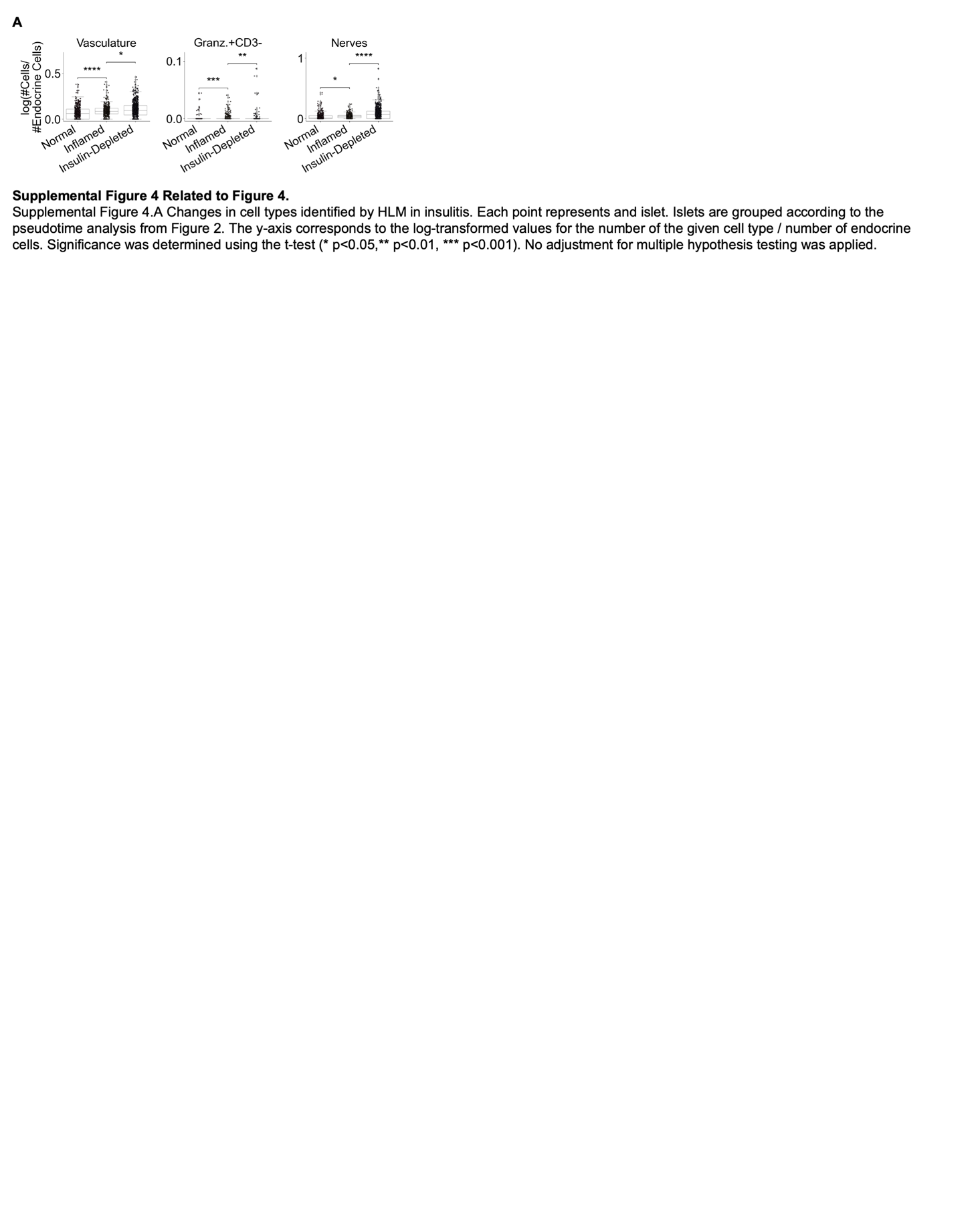

### Supplemental Figure 4 Related to Figure 4

Supplemental Figure 4 Changes in cell types identified by HLM in insulitis. Each point represents and islet. Islets are grouped according to the pseudotime analysis from Figure 2. The y-axis corresponds to the log-transformed values for the number of the given cell type / number of endocrine cells. Significance was determined using the t-test (* p<0.05,** p<0.01, *** p<0.001). No adjustment for multiple hypothesis testing was applied.

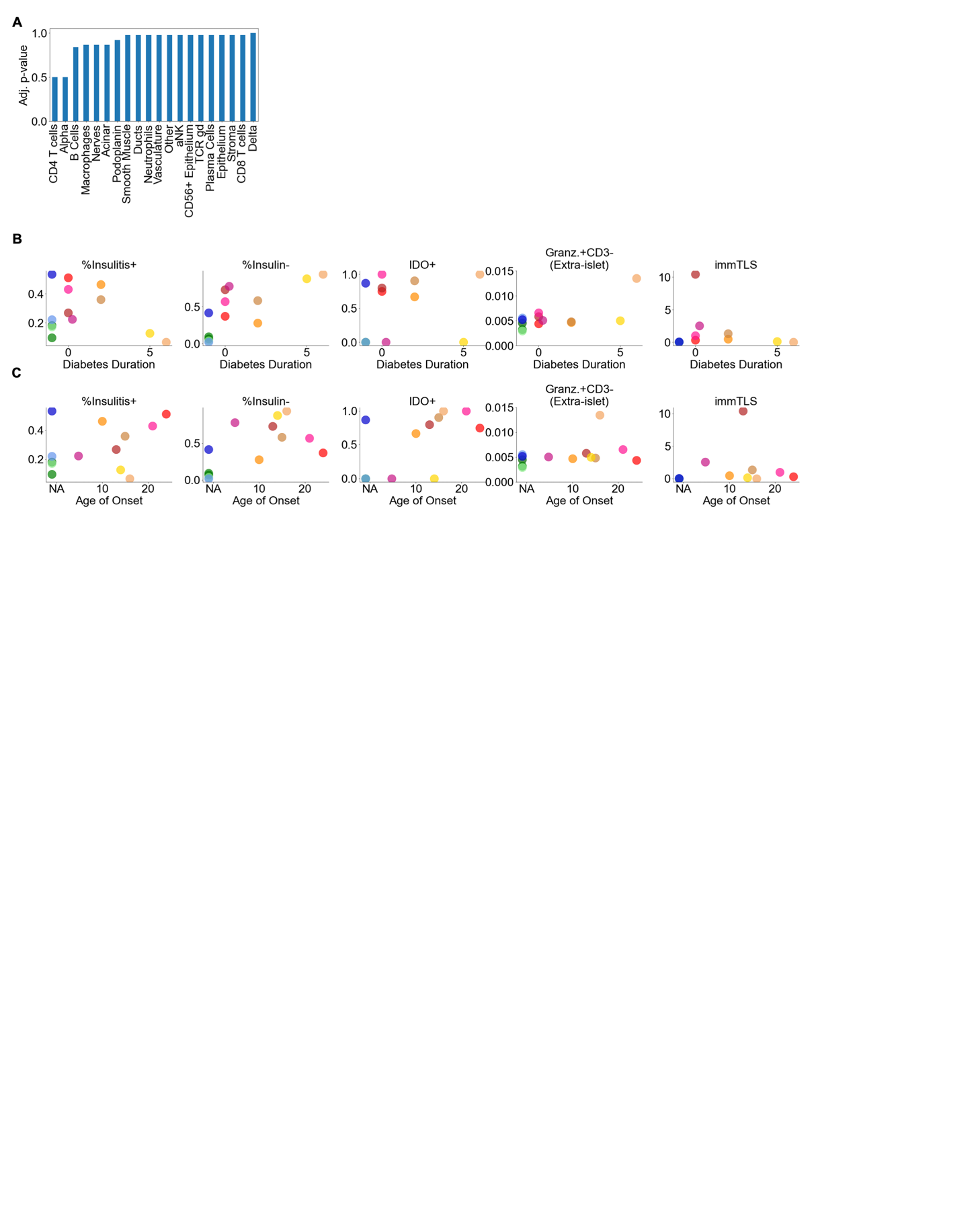

### Supplemental Figure 5 Related to Figure 5

Supplemental Figure 5.A Association of the cell composition of the CD8^+^T cells|B Cells CN with islet proximity. For each feature (columns), a mixed-level model adjusting for donor was fit against instances that were or were not < 20µm from an islet and the p-value was determined using Satterthwaites’s method in lmerTest R package. Values in the heatmap were corrected for multiple hypotheses using the Benjamini, Hochberg method.

Supplemental Figure 5.B Correlation of key islet features with diabetes duration.

Supplemental Figure 5.C Correlation of key islet features with age of onset.

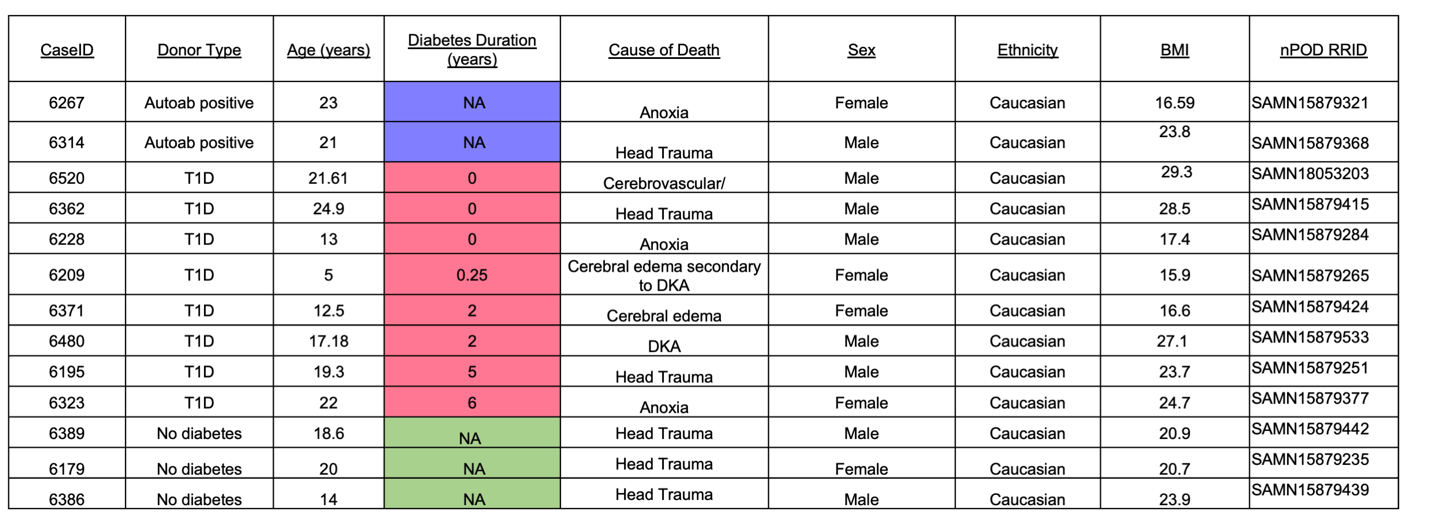

**Supplemental Table 1**: **nPOD Case Information**

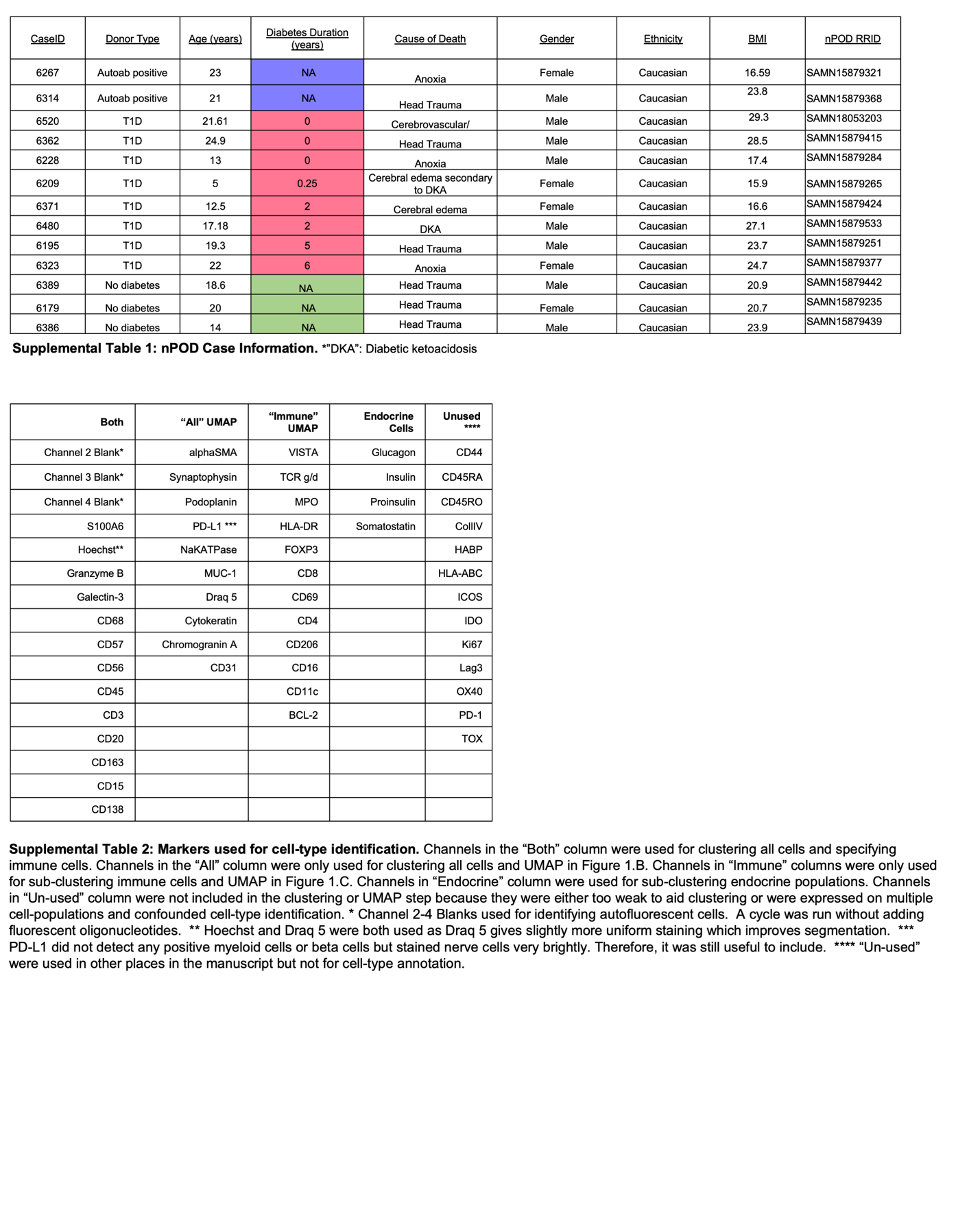

**Supplemental Table 2**: **Markers used for cell-type identification.** Channels in the “Both” column were used for clustering all cells and specifying immune cells. Channels in the “All” column were only used for clustering all cells and the UMAP in Figure 1.B. Channels in “Immune” columns were only used for sub-clustering immune cells and the UMAP in Figure 1.C. Channels in “Endocrine” column were used for sub-clustering endocrine populations. Channels in “Un-used” column were not included in the clustering or UMAP step because they were either too weak to aid clustering or were expressed on multiple cell-populations and confounded cell-type identification.

*Main Panel*

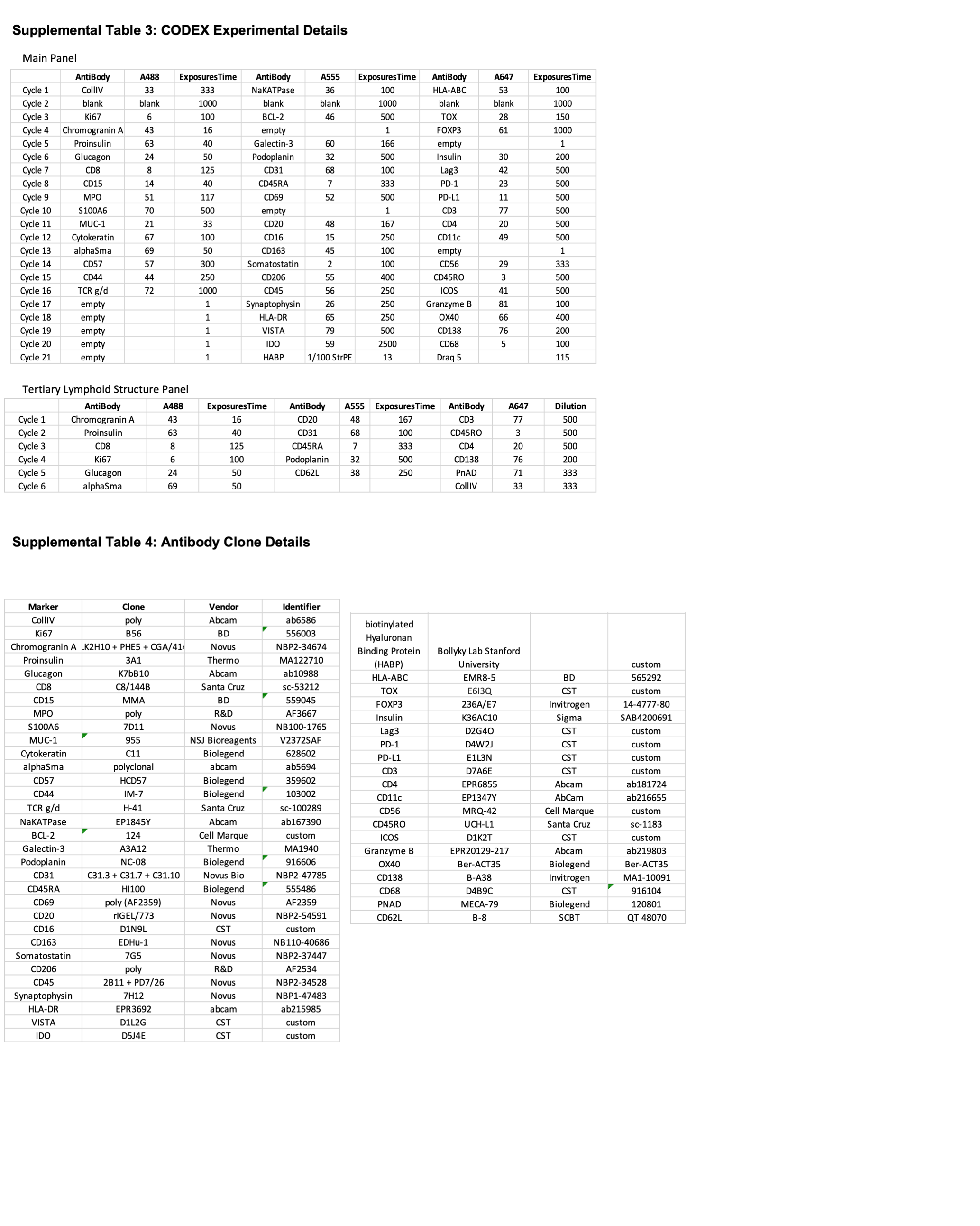

*Tertiary Lymphoid Structure Panel*

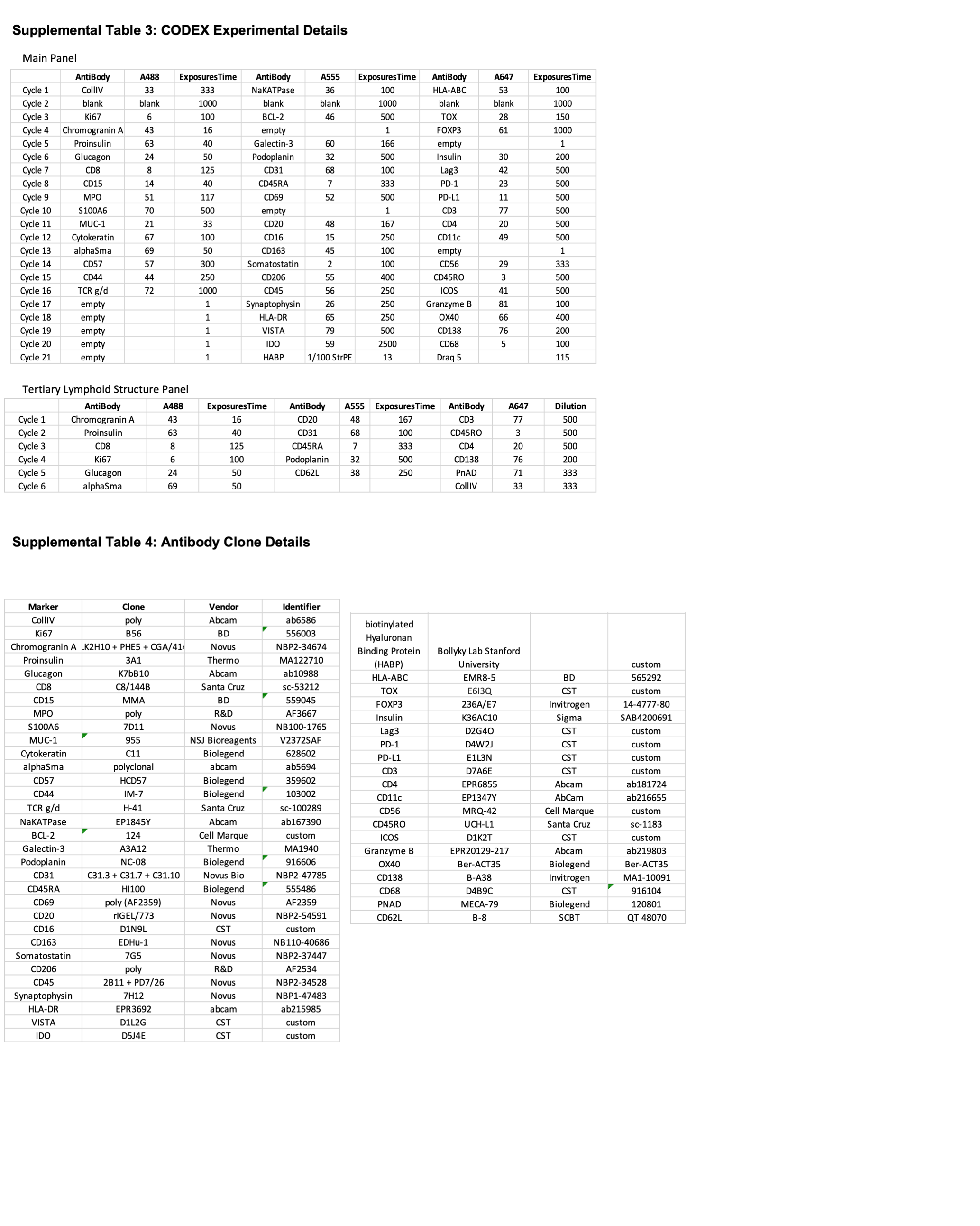

**Supplemental Table 3: CODEX Experiment Details**

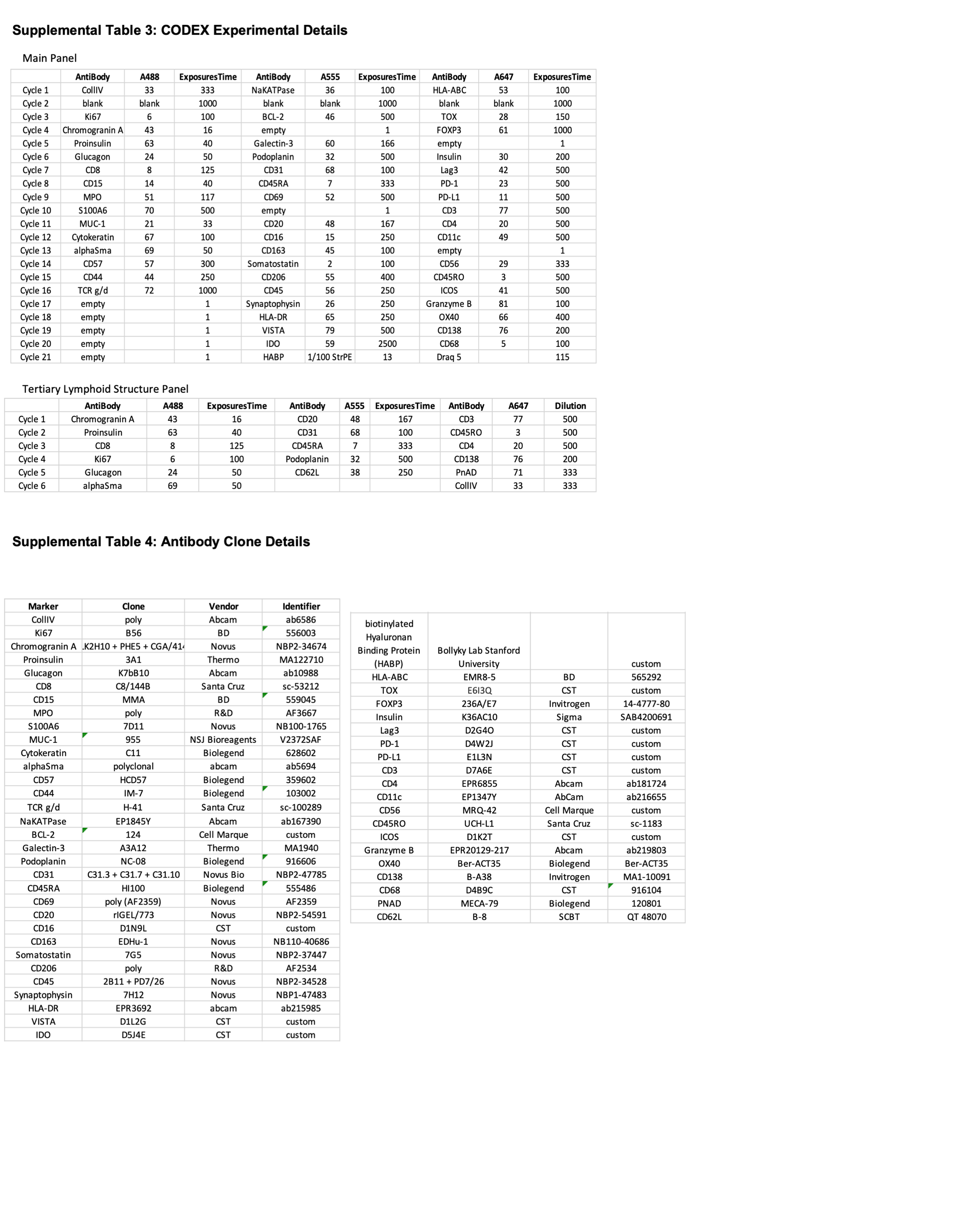

**Supplemental Table 4**: **Antibody Clone Details**
